# Associations between Maternal Sucrose-stimulated Salivary pH and Early Childhood Caries Diagnosis and Severity: An Observational Study

**DOI:** 10.1101/2023.10.30.23297786

**Authors:** David Okuji, Amy Lin, Olutayo Odusanwo, Yinxiang Wu

**Affiliations:** Senior Associate Director, NYU Langone Dental Medicine, Hansjörg Wyss Department of Plastic Surgery, Division of Dental Medicine, NYU Grossman School of Medicine, 5800 Third Avenue, 3rd Floor, Brooklyn, NY 11220; Pre-doctoral student, Harvard School of Dental Medicine, Harvard University; Faculty, Advanced Education in Pediatric Dentistry, NYU Langone Dental Medicine, Hansjörg Wyss Department of Plastic Surgery, Division of Dental Medicine, NYU Grossman School of Medicine and Private Practice, GA; Former Assistant Research Scientist, NYU Langone Health, Division of Biostatistics, Department of Population Health

**Author notes:** CORRESPONDING AUTHOR: David Okuji, DDS, MBA, MS NYU Langone Hospitals 5800 Third Avenue, 3^rd^ Floor Brooklyn, NY 11220.

**Keywords:** dentists, maternal, sucrose, salivary-pH, early-childhood-caries

## Abstract

**Background:** Multiple studies indicate that maternal sucrose-stimulated salivary pH has the potential to signal higher risk for diagnoses of early childhood caries and severity categories from the vertical transmission of bacteria from mother to child.

**Aim:** This study primarily investigated the relationship between maternal sucrose-stimulated salivary pH and child caries diagnosis and severity. It secondarily validated associations between 1) child and maternal sucrose-stimulated salivary pH, and 2) child sucrose-stimulated salivary pH and caries diagnosis and severity.

**Design:** Salivary pH levels were measured from 440 mother-child dyads. Early childhood caries diagnoses and severity levels were recorded. The analysis of variance test identified the associations between child and maternal sucrose-stimulated salivary pH and regression models were derived to show that maternal and child salivary pH were estimators for children’s caries diagnosis and severity.

**Results:** Maternal salivary pH <= 5.6, compared to maternal salivary pH = 7, had 7.58 times higher odds for the child’s ECC diagnosis and 5.61 times higher odds for moderate-extensive caries severity. Similar results were found for child salivary pH <= 5.6, compared to child salivary pH = 7. Maternal and child salivary pH were positively associated. When maternal salivary pH, compared to child salivary pH, was included with age, gender, and race/ethnicity as predictors for early childhood caries, the median interquartile ranges for sensitivity increased from 58.8 to 64.7 percent and specificity decreased from 84.6 to 80.8 percent.

**Conclusions:** Dentists should understand that both maternal and child sucrose-stimulated salivary pH can signal the diagnosis and severity of early childhood caries.

## INTRODUCTION

Dental caries is the most common chronic condition inflicting children, and if left untreated, poses a significant cost on society.^1^ Guidelines have been established by American Academy of Pediatric Dentistry (**AAPD**) and the International Caries Detection and Assessment System (**ICDAS**) on how each, respectively, define early childhood caries and caries severity.

The AAPD defines early childhood caries (**ECC**) as the presence of one or more decayed primary tooth surface in a child less than or equal to 71 months.^2^ The ICDAS system classifies caries severity with seven levels ranging from zero (sound tooth surface) to six (extensive enamel cavity).^3^

Children with ECC miss more days of school, have increased inappropriate use of over-the-counter pain medications, and experience diminished quality of life. Low maternal education, low family SES, and increased dietary refined carbohydrates have been established as conventional risk factors for ECC.^4,5^ Despite numerous advancements that have been made in understanding the biology and mechanisms behind the development of caries, current ECC risk assessment tools have shown limited efficacy in predicting caries risk.^6^

Saliva plays a pivotal role in maintaining oral homeostasis, as it protects the oral mucosa and assists with the neutralization of acids to promote the remineralization of teeth.^7^ It provides a protective layer on the tooth surface with the help of calcium, phosphate, and proteins that act as antibacterial substances and buffers. However, salivary composition can be altered by systemic conditions. The *Mutans streptococci* (***MS***) group of bacteria includes *Streptococci mutans (**S. mutans**)* and *Streptococci sobrinus* (***S. sobrinus***) which are, respectively, responsible for caries initiation and progressive development.^8–10^ Early acquisition of MS and higher levels of salivary MS in infants have been shown to be a major risk factor for ECC and future caries experience.^11,12^

Previous research has shown an association between child salivary pH and early childhood caries.^13,14^ The quantity of microorganisms influences salivary pH, which in turn impact the buffering capacity of saliva.^13^ Low salivary pH has been shown to be positively correlated with ECC and increases the odds of more extensive caries severity.^14^ Also, maternal and child salivary pH have been found to be positively associated.^14^

During pregnancy, human saliva composition changes with variations in female steroid sex hormones which alter the oral microbiome. It has been found that pregnant women have a lower salivary pH, which has been associated with increased risk of caries due to the microflora influences of cariogenic *S. mutans* and *Lactobacilli*.^10,13,15–20^ There is an increase in the decayed missing filled teeth (**DMFT**) index throughout pregnancy.^21^

During early childhood, initial MS colonization in infants occurs between seven and 36 months, coinciding with the timepoint of primary tooth eruption.^10,22^ *S. mutans* metabolizes carbohydrates to produce acid, further enhancing its cariogenic properties through maintenance of an acidic pH (<5.5) environment.^10^

It has been shown that there exists a strong link between maternal and infant oral microbiome and health. Vertical transmission of bacteria from mother to child occurs through saliva, and transmission is influenced by the level of bacteria contained in maternal saliva, frequency of contact between maternal and child’s saliva, age, and child’s salivary flow and diet.^10,23–25^ MS is transmitted from the mother to her infant, and there is increased risk of infant acquisition if the mother has high salivary levels of MS.^10^ Mothers of children with ECC have a higher average DMFT index,^23^ and poor maternal oral health has been associated with poor oral hygiene and ECC in their children.^23,26,27^ MS from the infant and their mother’s salivary samples share identical bacteriocin profiles and plasmid or chromosomal DNA patterns, and very little MS genotype commonality existed between the infants and their fathers.^10,11,25,28^ Reduction of maternal MS during the emergence of primary teeth in her child has a long-term influence on the child’s oral bacterial colonization and caries experience, preventing or delaying colonization of these bacteria in the child and thus caries development.^29^

Given the vertical transmission of cariogenic bacteria, the link between maternal and infant oral microflora, and evidence that maternal sucrose-stimulated salivary pH (**SSSpH**) is associated with child SSSpH, the primary purpose of this study was to determine if there is an association between maternal SSSpH and early childhood caries diagnosis and severity in their children, with secondary objectives to validate the association between child- and maternal- SSSpH and the association between child SSSpH and ECC diagnosis and severity. The null hypothesis for the primary question is that there is no association between maternal SSSpH and child ECC diagnosis and severity. The null hypotheses for the secondary questions are that there are no associations between maternal and child SSSpH and there are also none between child SSSpH and child ECC diagnosis and severity.

## METHODS

### Study design and modeling

An observational diagnostic study design was conducted to collect and measure SSSpH from mothers and their children and classify the children’s caries diagnosis and severity, respectively, with the AAPD definition of early childhood caries and the ICDAS classification system for caries activity and severity.

The study was modeled on one of the outcomes from the classic Stephan experiment which found that after oral exposure to simple carbohydrates, the pH level of saliva and plaque decrease and gradually return to resting saliva and plaque pH level within thirty minutes after exposure.^4,30,31^ Children and their biologic mothers who met the study’s inclusion criteria were exposed to oral sucrose and their salivary pH was measured thirty minutes after exposure. The study was also modeled on the principles of implementation science, which is defined as “the study of methods to promote the adoption and integration of evidence-based practices, interventions and policies into routine health care and public health settings.^32^” The study design implemented evidence-based science into clinical practice for one component of the classic Stephan experiment as a cost- and time-effective screening tool to identify children for caries diagnosis and severity.

### Study sample size and setting

A power analysis based upon a previous study’s results,^14^ for the association between maternal and child SSSpH, yielded a power level over 80% at alpha equal to 0.05, with a sample size 119 to demonstrate a sufficient statistical power level.

The study was approved by the NYU Grossman School of Medicine Institutional Review Board under protocol number 18-00076. Pediatric subjects and their biologic mothers were selected from the pool of consecutive mother-child dyads presenting to the dental clinic between February 2019 and December 2020, who met the inclusion criteria at NYU Langone Hospitals-affiliated training sites located in Hawaii, New York, and Tennessee.

Mothers and their biologic children were included in the sample if the children were 0-5 years in age, categorized as American Society of Anesthesiologists (**ASA**) Physical Status Classification System of 1 (normal, healthy patient) or 2 (patient with mild systemic disease),^33^ were a patient of record at an NYU Langone Hospitals-affiliated training site, had at least one erupted tooth, had a caries risk and had an oral evaluation for ECC diagnosis and ICDAS classification.

### Data collection and Quantiztative Variables

Resident-researchers enrolled in the NYU Langone Hospitals-Advanced Education in Pediatric Dentistry residency program performed data collection after attending a training conference to ensure one standardized protocol across three study locations. Following the sample selection, corresponding clinical charts were reviewed using electronic medical record software. Biologic mothers of pediatric subjects, who met the inclusion criteria, were invited to participate in the study. At the time of the visit, mothers provided informed written consent to participate in the study. Unlike a previous study^14^ which instructed subjects to not brush their teeth for 12-hours prior to salivary sampling, this study instructed the mother- and child-subjects to not brush their teeth at least 1-hour prior to the sucrose challenge at the next visit. The reason for this change is because in a previous study,^14^ many of the mother-subjects declined to participate in the study because they did not want to defer toothbrushing for 12-hours prior to salivary sampling. Based upon the stages of oral biofilm development, this present study’s premise was that there may be differences in SSSpH for subjects who last brushed their teeth less than 3-hours, during the initial colonization of the oral biofilm, compared to those who last brushed their teeth greater than or equal to 12-hours, during the rapid growth and extracellular polysaccharide production stage of the oral biofilm.^34^

At the next visit, one packet of table sugar (sucrose) was administered to the biologic mother and child subjects by dissolving the sugar granules on the mother’s and child’s tongue, respectively, and mother and child were asked to wait approximately 30 minutes before collection of their respective saliva specimen. During the 30-minute waiting period, the mother was asked to complete a sociodemographic questionnaire. After the sucrose challenge, the biologic mother and pediatric subjects were instructed to provide a stimulated saliva specimen into a cup. Study participants expectorated whole saliva into a cup for about 1-minute. The resident-researcher measured the pH levels of the biologic mother’s and child’s salivary specimens by dipping pH strips into the provided saliva specimen and compared immediate color change with a chart provided by the manufacturer (Precision pH 4070 test strips, Precision Laboratories, Cottonwood, AZ) and recorded the child’s and mother’s pH level, quantified by the manufacturer’s strip color chart, into the child subject’s electronic dental record. Any residual saliva specimen was discarded in a sink, followed by flushing with water. Clinical data from the dental record, including the child’s caries diagnosis and caries severity (using the merged-ICDAS system), were abstracted after the collection of saliva specimens.^31,35,36^

The quantitative variables were defined as listed below:

? SSSpH: Measured in eight ordinal increments at pH levels 4.0, 4.4, 4.8, 5.2, 5.6, 6.0, 6.5, and 7.0.
? ECC: Defined as “the presence of one or more decayed (noncavitated or cavitated lesions), missing (due to caries), or filled tooth surfaces in any primary tooth in a child under the age of six^2^”, as diagnosed by a dentist-provider.
? SECC: Defined as “1) any sign of smooth-surface caries in a child younger than three years of age, 2) from ages three through five, one or more cavitated, missing (due to caries), or filled smooth surfaces in primary maxillary anterior teeth, or 3) a decayed, missing, or filled score of greater than or equal to four (age three), greater than or equal to five (age four), or greater than or equal to six (age five) ^2^” as diagnosed by a dentist- provider.
? ICDAS Categories^3^: Defined as:

- Zero = Sound tooth surface: No evidence of caries after 5 sec air drying.
- 1 = First visual change in enamel: Opacity or discoloration (white or brown) is visible at the entrance to the pit or fissure seen after prolonged air drying.
- 2 = Distinct visual change in enamel visible when wet, lesion must be visible when dry.
- 3 = Localized enamel breakdown (without clinical visual signs of dentinal involvement) seen when wet and after prolonged drying.
- 4 = Underlying dark shadow from dentine.
- 5 = Distinct cavity with visible dentine.
- 6 = Extensive (more than half the surface) distinct cavity with visible dentine.
? Time elapsed since child and biologic mother last brushed their teeth defined with the response options as:

- no response
- < 1-hour
- >= 1 < 2-hours
- >= 2 < 3-hours
- >= 3 < 4-hours
- >= 4 < 5-hours
- >= 5 < 6-hours
- >= 6 < 7-hours
- >= 7 < 8-hours
- >= 8 < 9-hours
- >= 9 < 10-hours
- >= 10 < 11-hours
- >= 11 < 12-hours
- >= 12-hours

### Statistical analysis

Patient demographic and clinical characteristics were summarized in a descriptive table using mean and standard deviations for continuous variables and using counts and percentages for categorical variables. The distributions of variables were compared across groups defined by child caries diagnosis. When assessing the correlation between child and mother salivary pH, the Kendall’s tau rank correlation test was utilized. When analyzing associations with child salivary pH and fitting further regression models, the original pH levels were categorized into four levels, 5.6 or below, 6, 6.5, and 7, due to the small number of child subjects with pH < 5.6.

The associations of child and mother salivary pH were investigated separately with different dental outcomes, including child caries diagnosis, ICDAS caries classification, and the number of teeth which presented with the greatest degree of ICDAS dental caries severity classification. For child caries diagnosis, a binary outcome, namely, ECC/SECC vs. no dental caries was considered. For merged ICDAS caries classification, the binary outcome, i.e., extensive decay/moderate decay vs. initial stage decay/sound enamel, was considered. The binary outcomes were analyzed with logistic regressions. For the number of teeth with the greatest degree of dental caries severity, linear regression models were fitted. All models controlled for child age, gender, race/ethnicity. Adjusted odds ratios were reported from fitted logistic/cumulative odds models, and beta coefficients were reported from fitted linear regression models. The likelihood ratio test was used to assess model goodness of fit.

Sensitivity and specificity were calculated based on cross-validation to assess predictability of child and mother salivary pH for ECC/SECC. The logistic model with child age, gender, race/ethnicity, and salivary pH as predictors, was used as the main prediction model. 10-fold cross validation was conducted. The sample with complete data of all predictors and outcomes was randomly partitioned into 10 equal sized subsamples. The random sampling was done within the levels of the outcome to balance the class distribution within the splits. Of the 10 subsamples, a single subsample is retained as the validation data for testing the prediction model and the remaining 9 subsamples were used to fit the model. The cross-validation process was repeated 10 times, with each of the 10 subsamples used exactly once as the validation data. The entire 10-fold cross-validation was repeated 10 times. Sensitivity and specificity estimates from 10*10 validation data sets were reported and the results were compared to that from the model with only child age, gender, race/ethnicity as the predictors.

All statistical analyses were performed in R version 4.0.3 for Mac OS.^37^ Significance level was set at 0.05.

## RESULTS

The analytical sample size was 440 subjects. Table 1 presents the overall descriptive distribution of child-level demographic and clinical characteristics, and dichotomous child and mother salivary pH. The pediatric subjects were 42.3 percent female and the average ± standard deviation (SD) age was 3.19 ± 1.38. The distribution of race and ethnicity was 60.5 percent non-Hispanic White, 20.0 percent Hispanic, 6.4 percent non-Hispanic Black, and 12.5 percent non-Hispanic other/multi-race. The majority (97.3 percent) of the subjects were healthy as defined by ASA classification. Notably, 21 percent and 19 percent were diagnosed with early childhood caries (**ECC**) and severe early childhood caries (**SECC**), respectively.

**Table 1.**
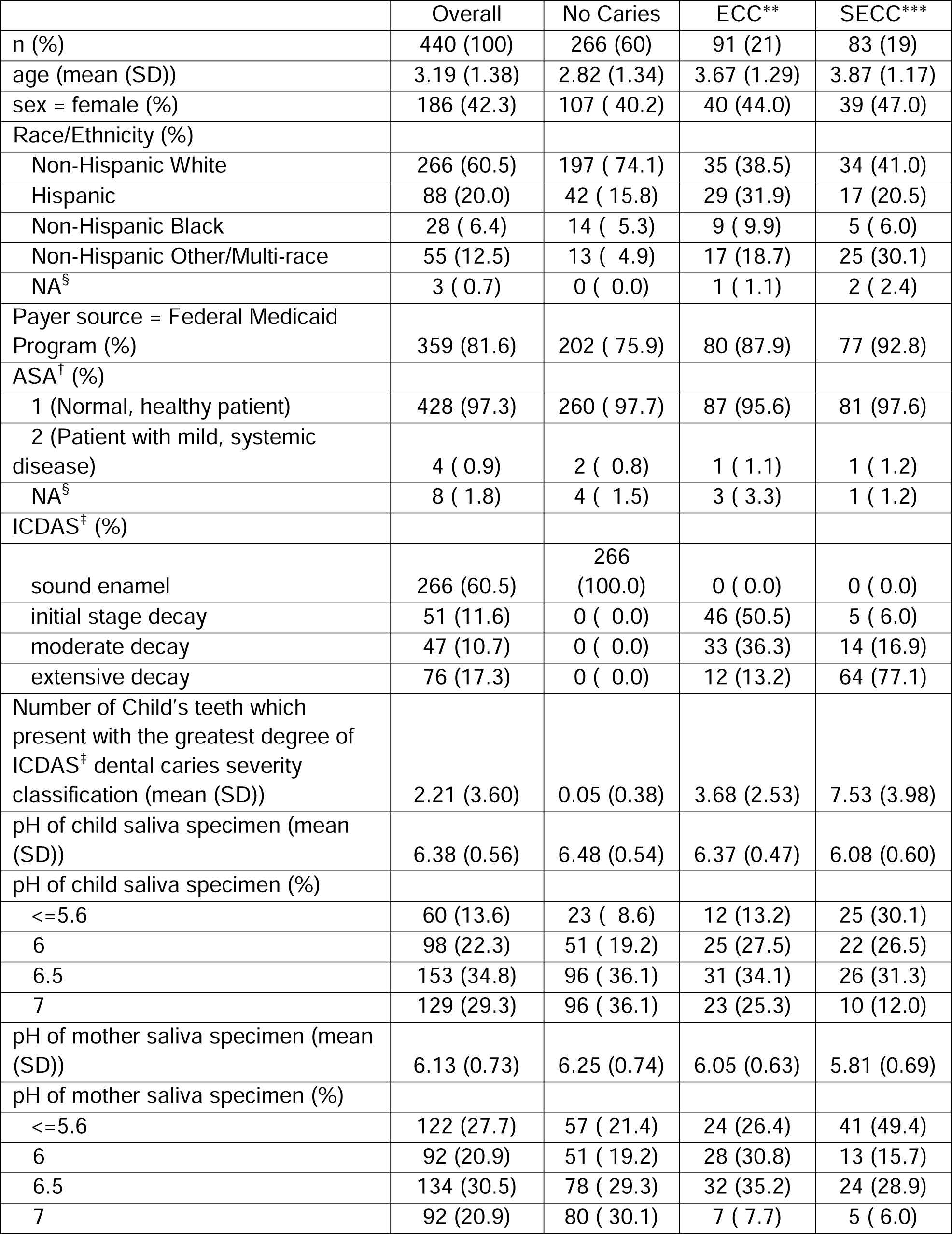
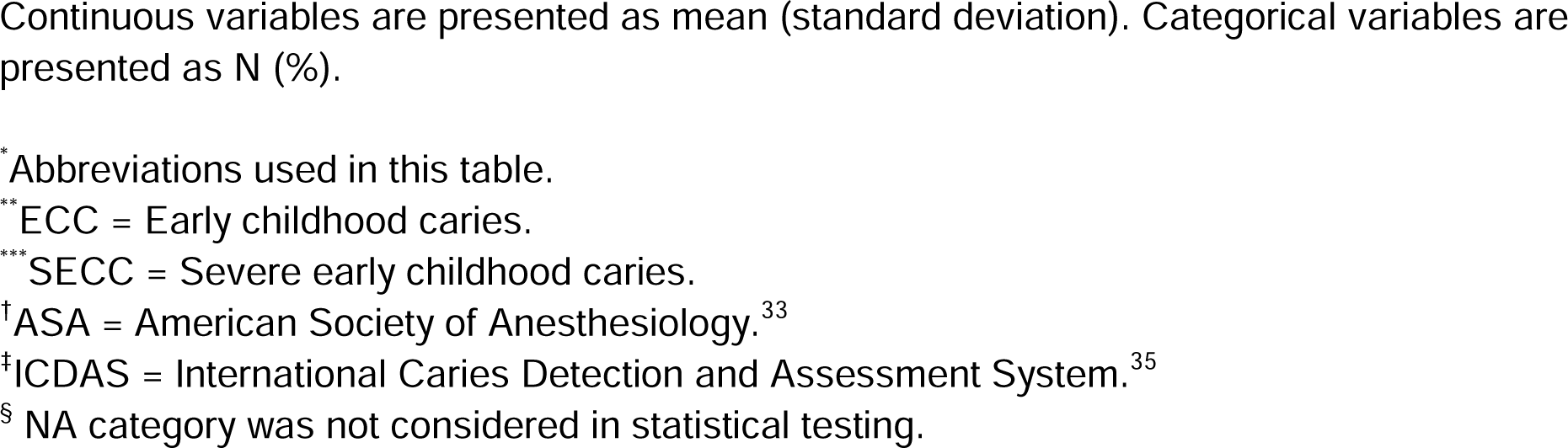
Demographic and clinical characteristics of child subjects, overall and by child diagnosed caries status. (N=440) *.

|  | Overall | No Caries | ECC** | SECC*** |
| --- | --- | --- | --- | --- |
| n (%) | 440 (100) | 266 (60) | 91 (21) | 83 (19) |
| age (mean (SD)) | 3.19 (1.38) | 2.82 (1.34) | 3.67 (1.29) | 3.87 (1.17) |
| sex = female (%) | 186 (42.3) | 107 ( 40.2) | 40 (44.0) | 39 (47.0) |
| Race/Ethnicity (%) |  |  |  |  |
| Non-Hispanic White | 266 (60.5) | 197 ( 74.1) | 35 (38.5) | 34 (41.0) |
| Hispanic | 88 (20.0) | 42 ( 15.8) | 29 (31.9) | 17 (20.5) |
| Non-Hispanic Black | 28 ( 6.4) | 14 ( 5.3) | 9 ( 9.9) | 5 ( 6.0) |
| Non-Hispanic Other/Multi-race | 55 (12.5) | 13 ( 4.9) | 17 (18.7) | 25 (30.1) |
| NA<sup>§</sup> | 3 ( 0.7) | 0 ( 0.0) | 1 ( 1.1) | 2 ( 2.4) |
| Payer source = Federal Medicaid Program (%) | 359 (81.6) | 202 ( 75.9) | 80 (87.9) | 77 (92.8) |
| ASA<sup>†</sup> (%) |  |  |  |  |
| 1 (Normal, healthy patient) | 428 (97.3) | 260 ( 97.7) | 87 (95.6) | 81 (97.6) |
| 2 (Patient with mild, systemic disease) | 4 ( 0.9) | 2 ( 0.8) | 1 ( 1.1) | 1 ( 1.2) |
| NA<sup>§</sup> | 8 ( 1.8) | 4 ( 1.5) | 3 ( 3.3) | 1 ( 1.2) |
| ICDAS<sup>‡</sup> (%) |  |  |  |  |
| sound enamel | 266 (60.5) | 266 (100.0) | 0 ( 0.0) | 0 ( 0.0) |
| initial stage decay | 51 (11.6) | 0 ( 0.0) | 46 (50.5) | 5 ( 6.0) |
| moderate decay | 47 (10.7) | 0 ( 0.0) | 33 (36.3) | 14 (16.9) |
| extensive decay | 76 (17.3) | 0 ( 0.0) | 12 (13.2) | 64 (77.1) |
| Number of Child's teeth which present with the greatest degree of ICDAS<sup>‡</sup> dental caries severity classification (mean (SD)) | 2.21 (3.60) | 0.05 (0.38) | 3.68 (2.53) | 7.53 (3.98) |
| pH of child saliva specimen (mean (SD)) | 6.38 (0.56) | 6.48 (0.54) | 6.37 (0.47) | 6.08 (0.60) |
| pH of child saliva specimen (%) |  |  |  |  |
| <=5.6 | 60 (13.6) | 23 ( 8.6) | 12 (13.2) | 25 (30.1) |
| 6 | 98 (22.3) | 51 ( 19.2) | 25 (27.5) | 22 (26.5) |
| 6.5 | 153 (34.8) | 96 ( 36.1) | 31 (34.1) | 26 (31.3) |
| 7 | 129 (29.3) | 96 ( 36.1) | 23 (25.3) | 10 (12.0) |
| pH of mother saliva specimen (mean (SD)) | 6.13 (0.73) | 6.25 (0.74) | 6.05 (0.63) | 5.81 (0.69) |
| pH of mother saliva specimen (%) |  |  |  |  |
| <=5.6 | 122 (27.7) | 57 ( 21.4) | 24 (26.4) | 41 (49.4) |
| 6 | 92 (20.9) | 51 ( 19.2) | 28 (30.8) | 13 (15.7) |
| 6.5 | 134 (30.5) | 78 ( 29.3) | 32 (35.2) | 24 (28.9) |
| 7 | 92 (20.9) | 80 ( 30.1) | 7 ( 7.7) | 5 ( 6.0) |

Continuous variables are presented as mean (standard deviation). Categorical variables are presented as N (%).
\* Abbreviations used in this table.
\*\* ECC = Early childhood caries.
\*\*\* SECC = Severe early childhood caries.
† ASA = American Society of Anesthesiology.<sup>33</sup>
‡ ICDAS = International Caries Detection and Assessment System.<sup>35</sup>
§ NA category was not considered in statistical testing.

The subjects diagnosed with ECC/SECC were on average older and with lower representation in the non-Hispanic White race/ethnicity sample. Both pH of child and mother saliva specimens were observed to be associated with child caries status (Table 1 and Figure 1). For the child-subjects with ECC or SECC, the child’s and their mother’s SSSpH were lower than those with no dental caries (Figure 1: (A) and (C)). Similar patterns can be found with ICDAS (Figure 1: (B) and (D)). Table 2 also shows that for both the children and mothers who had longer durations of time elapsed since their last toothbrushing, compared to those with lower durations, presented with higher levels of ECC/SECC diagnoses and higher severity of decay for the children.

**Figure 1.**
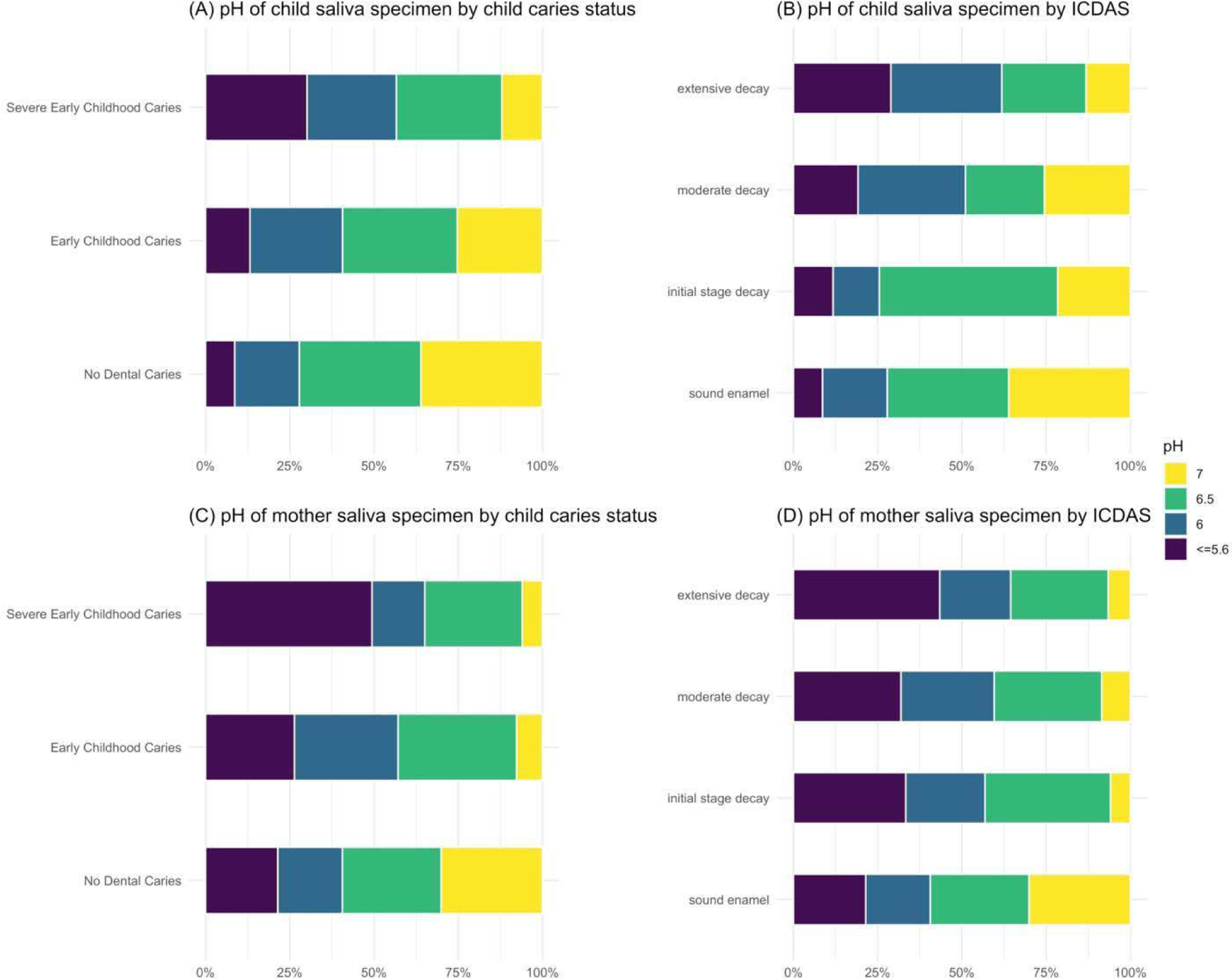
Percentage stacked bar charts by diagnosed child caries and merged-ICDAS caries classification. (A-B): pH levels of child saliva by diagnosed child caries and merged-ICDAS caries classification; (C-D): pH levels of mother saliva by diagnosed child caries and merged-ICDAS caries classification. Yellow bars correspond to pH level = 7; Dark blue bars correspond to pH level = 5.6 or less.

**Table 2.** Time elapsed since last tooth brushing of child- and maternal-subjects, overall and by 1) child caries diagnosis, 2) caries severity status, and 3) child salivary pH. *.

|  | **Overall** | **No caries** | **ECC\*\* and SECC\*\*\*** |
| --- | --- | --- | --- |
| n | 440 | 262 | 178 |
| Time elapsed since CHILD last had his/her teeth brushed (hours) (mean (SD)) | 7.29 (3.91) | 6.78 (4.15) | 8.04 (3.56) |
| Time elapsed since biologic mother last had brushed her teeth (hours) (mean (SD)) | 7.22 (3.79) | 6.70 (3.99) | 7.98 (3.48) |
|  | **Overall** | **Sound enamel** | **Initial stage decay, moderate decay, and extensive decay** |
| n | 440 | 265 | 175 |
| Time elapsed since CHILD last had his/her teeth brushed (hours) (mean (SD)) | 7.27 (3.93) | 6.83 (4.15) | 8.09 (3.39) |
| Time elapsed since biologic mother last had brushed her teeth (hours) (mean (SD)) | 7.27 (3.77) | 6.75 (3.99) | 8.01(3.33) |
|  | **Overall** | **Child salivary pH ≤ 5.6** | **Child salivary pH > 5.6** |
| n | 440 | 60 | 380 |
| Time elapsed since CHILD last had his/her teeth brushed (hours) (mean (SD)) | 7.17 (4.01) | 7.24 (3.73) | 7.17 (4.05) |
| Time elapsed since biologic mother last had brushed her teeth (hours) (mean (SD)) | 7.10 (3.88) | 6.97 (3.41) | 7.11 (3.94) |

\*Abbreviations used in this table.
\*\*ECC = Early childhood caries.
\*\*\*SECC = Severe early childhood caries.

Table 3 presents descriptive differences in demographic and clinical characteristics by the pH of child saliva specimen. Except for race/ethnicity, no associations with demographics were observed. The pH of child and mother saliva were observed to be positively associated with a low magnitude Kendall’s tau rank correlation = 0.23 (p<0.001). The child subjects with salivary pH less than or equal to 6.0 had greater tendency to have overall moderate/extensive decay on teeth, ECC/SECC, and more teeth with greatest degree of caries severity. Table 2 also shows that for both the children and mothers, there were little differences between the duration of time elapsed since their last toothbrushing and the child’s SSSpH.

**Table 3.** Demographic and clinical characteristics of child subjects, overall and by saliva pH levels of child subjects. *.

|  | pH of child saliva specimen |  |  |  |
| --- | --- | --- | --- | --- |
|  | <=5.6 | 6 | 6.5 | 7 |
| n | 60 | 98 | 153 | 129 |
| age (mean (SD)) | 3.18 (1.43) | 3.24 (1.36) | 3.07 (1.39) | 3.30 (1.35) |
| sex = female (%) | 24 (40.0) | 38 (38.8) | 68 (44.4) | 56 (43.4) |
| Race/Ethnicity (%) |  |  |  |  |
| Non-Hispanic White | 23 (38.3) | 49 (50.0) | 91 (59.5) | 103 (79.8) |
| Hispanic | 19 (31.7) | 20 (20.4) | 40 (26.1) | 9 ( 7.0) |
| Non-Hispanic Black | 2 ( 3.3) | 4 ( 4.1) | 12 ( 7.8) | 10 ( 7.8) |
| Non-Hispanic Other/Multi-race | 14 (23.3) | 25 (25.5) | 10 ( 6.5) | 6 ( 4.7) |
| NA<sup>§§</sup> | 2 ( 3.3) | 0 ( 0.0) | 0 ( 0.0) | 1 ( 0.8) |
| Payor source = Federal Medicaid Program (%) | 54 (90.0) | 84 (85.7) | 122 (79.7) | 99 (76.7) |
| ASA<sup>†</sup> (%) |  |  |  |  |
| 1 (Normal, healthy patient) | 57 (95.0) | 94 (95.9) | 149 (97.4) | 128 (99.2) |
| 2 (Patient with mild, systemic disease) | 1 ( 1.7) | 0 ( 0.0) | 2 ( 1.3) | 1 ( 0.8) |
| NA<sup>§§</sup> | 2 ( 3.3) | 4 ( 4.1) | 2 ( 1.3) | 0 ( 0.0) |
| ICDAS<sup>‡</sup> (%) |  |  |  |  |
| sound enamel | 23 (38.3) | 51 (52.0) | 96 (62.7) | 96 (74.4) |
| initial stage decay | 6 (10.0) | 7 ( 7.1) | 27 (17.6) | 11 ( 8.5) |
| moderate decay | 9 (15.0) | 15 (15.3) | 11 ( 7.2) | 12 ( 9.3) |
| extensive decay | 22 (36.7) | 25 (25.5) | 19 (12.4) | 10 ( 7.8) |
| Number of Child's teeth which present with the greatest degree of ICDAS<sup>‡</sup> dental caries severity classification (mean (SD)) | 4.52 (4.64) | 2.94 (4.48) | 1.68 (2.63) | 1.22 (2.64) |
| Caries diagnosis = ECC<sup>**</sup>/SECC<sup>***</sup> (%) | 37 (61.7) | 47 (48.0) | 57 (37.3) | 33 (25.6) |
| pH of mother saliva specimen<sup>§</sup> (mean (SD)) | 5.75 (0.69) | 6.01 (0.76) | 6.19 (0.63) | 6.32 (0.74) |

Continuous variables are presented as mean (standard deviation). Categorical variables are presented as N (%).
\* Abbreviations used in this table.
\*\* ECC = Early childhood caries.
\*\*\* SECC = Severe early childhood caries.
† ASA = American Society of Anesthesiology.<sup>33</sup>
‡ ICDAS = International Caries Detection and Assessment System.<sup>35</sup>
§ p<0.001 from Kendall's tau rank test; Kendall's tau rank correlation = 0.23.
§§ NA category was not considered in statistical testing.

The adjusted associations of the pH of child or mother saliva specimen with different dental outcomes can be found in Table 4. Statistically significant associations of both maternal and child salivary pH were observed with all dental outcomes. Compared to those with salivary pH = 7 (or with child’s salivary pH = 7), maternal subjects with lower salivary pH (or lower child’s salivary pH) had greater odds of having ECC/SECC, severe decay on teeth, and an increased number of teeth with greatest severity of decay. In Model 1, the odds of child diagnosis of ECC/SECC for maternal subjects with salivary pH <= 5.6 were 7.58 [3.54, 17.45] times higher than that for mothers with salivary pH = 7. Similarly, child subjects with salivary pH <= 5.6 had 3.80 [1.76, 8.36] times higher odds of having ECC/SECC, compared to those with child salivary pH = 7. In Model 2, the odds of child caries severity of extensive/moderate decay for maternal subjects with salivary pH <= 5.6 were 5.61 [2.43, 14.40] times higher than that for mothers with salivary pH =7, with similar increased odds for child SSSpH at 4.10 [1.83, 9.39] compared to the child SSSpH 7.0 reference. In Model 3, the number of teeth with the highest ICDAS classification in children had 1.73 [0.88, 2.58] higher odds for maternal subjects with salivary pH <= 5.6 compared to mothers with salivary pH = 7, with similar increased odds for child SSSpH at 2.39 [1.41, 3.38] increased odds compared to the child SSSpH 7.0 reference.

**Table 4.**
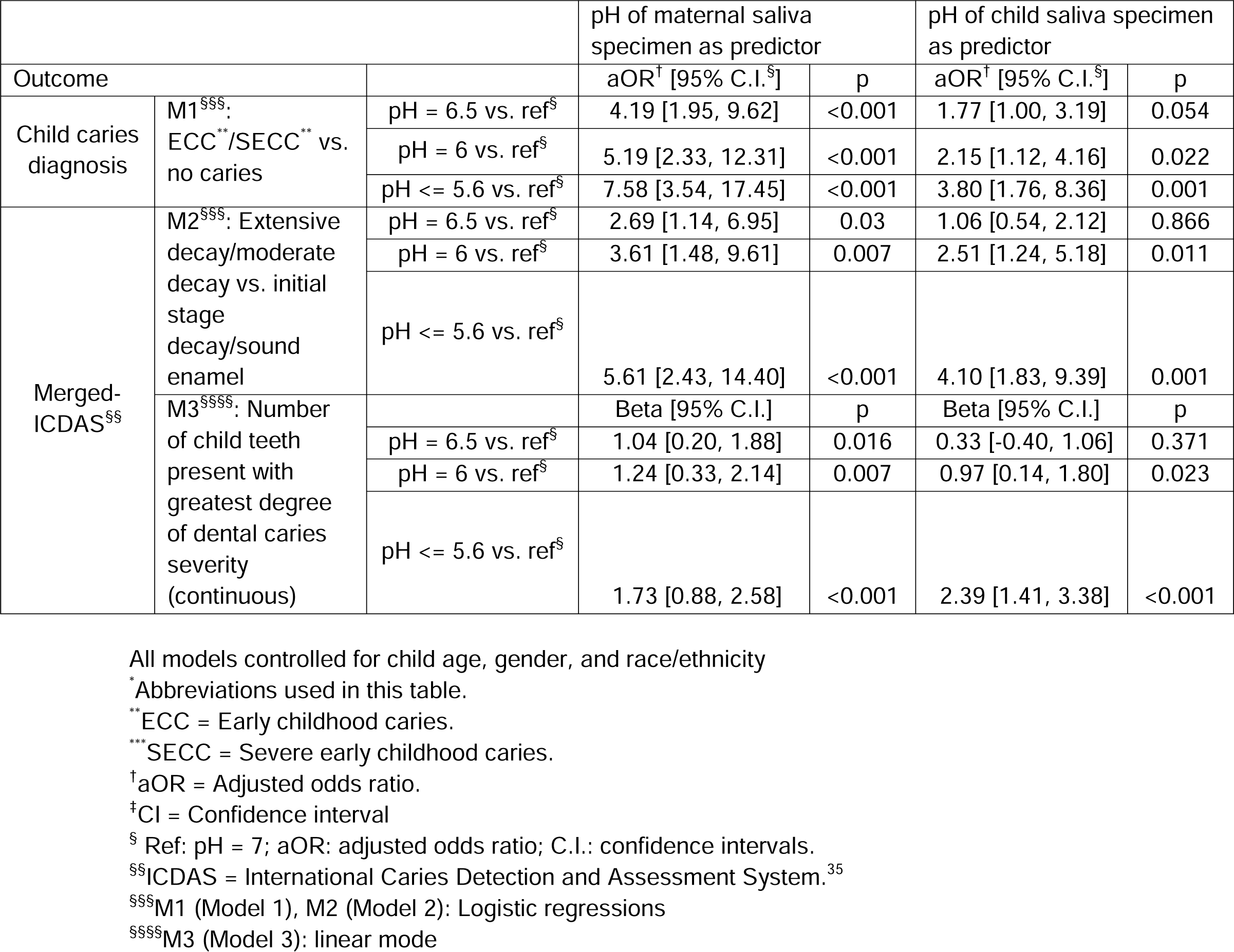
Associations of saliva pH levels with different dental outcomes. Results from logistic, cumulative odds, and regular linear models. Associations of child and mother salivary pH with different outcomes were estimated in separate models. Complete data N = 437 due to missingness in race/ethnicity.*

Finally, as visualized in Figure 2, when child age, gender, race/ethnicity, and child salivary pH were used as the predictors for ECC/SECC, the median [IQR: interquartile range] of sensitivity and specificity estimates from repeated 10-fold cross validation were respectively, 58.8 percent [47.1 percent, 64.7 percent] and 84.6 percent [76.9 percent, 88.5 percent]. If maternal salivary pH is used in the model to replace child salivary pH, the median [IQR] of sensitivity increased to 64.7 percent [52.9 percent, 70.6 percent], but the median [IQR] of specificity decreased to 80.8 percent [76.9 percent, 85.2 percent]. For comparison, the median [IQR] of sensitivity and specificity were respectively 47.1 percent [41.2 percent, 58.8 percent] and 85.2 percent [80.0 percent,88.9 percent] from the model with only child age, gender, race/ethnicity as the predictors.

**Figure 2.**
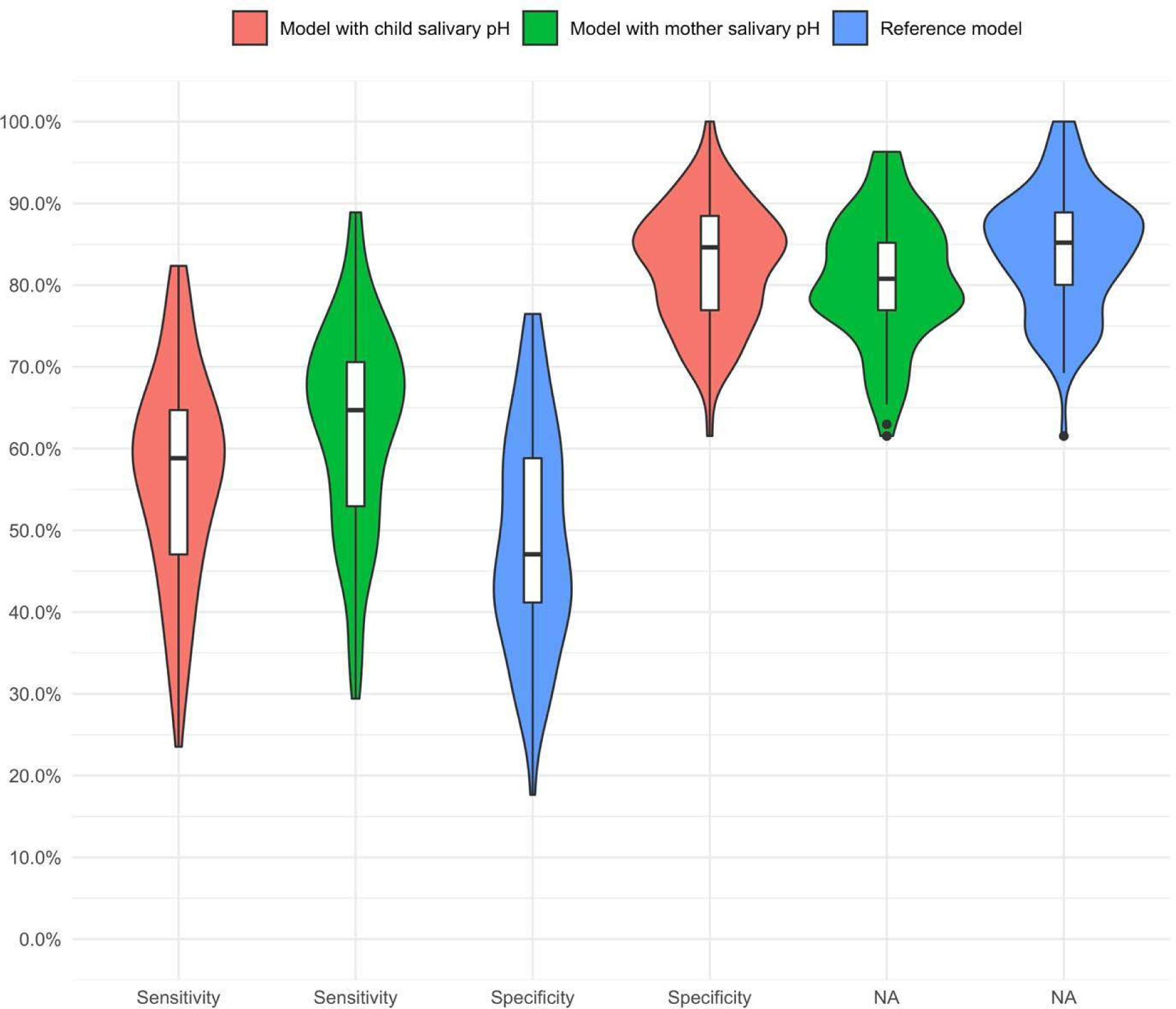
Violin plots for sensitivity and specificity estimates for predicting SECC/ECC from repeated 10-fold cross-validation. Sensitivity and specificity were calculated based on cross- validation to assess predictability of child salivary pH for SECC/ECC. The sample with complete data of all predictors and outcomes was N=437 and randomly partitioned into 10 equal sized subsamples. Of the 10 subsamples, a single subsample is retained as the validation data for testing the prediction model and the remaining 9 subsamples were used to fit the model. The cross-validation process was repeated 10 times, with each of the 10 subsamples used exactly once as the validation data. The random sampling was done within the levels of the outcome to balance the class distribution within the splits. The whole 5-fold cross-validation was repeated 10 times. Sensitivity and specificity of predicting SECC/ECC over 100 (10*10) validation data sets were reported. A violin plot with a box plot inside presents the distribution of 100 sensitivity/specificity estimates. The violin plots with red filled area correspond to the logistic model with child age, gender, race/ethnicity, and child salivary pH as the predictors. The violin plots with green filled area correspond to the logistic model with child age, gender, race/ethnicity, and mother salivary pH as the predictors. The violin plots with blue filled area correspond to the logistic model (reference model) with only child age, gender, and race/ethnicity as the predictors.

## DISCUSSION

A systematic search of the literature yielded a large body of evidence which supports the association of maternal risk factors with ECC. However, the search did not find evidence which focused on the association of maternal SSSpH with ECC and ICDAS caries severity. Therefore, this study’s primary research question may represent the first known investigation to report the association of maternal SSSpH with the diagnosis of ECC and ICDAS severity of caries classification. This study further validates previous reports for the associations for the secondary research questions of 1) maternal SSSpH with child SSSpH and 2) child SSSpH with the ECC diagnosis and severity. Hence, these results reject the null and accept the alternative hypothesis that there are statistically significant differences for the associations between 1) lower maternal SSSpH and ECC diagnosis and severity, 2) lower maternal SSSpH with lower child SSSpH, and 3) lower child SSSpH and ECC diagnosis and severity.

Studies which support the association of maternal risk factors with early childhood caries show that mothers of children with ECC have a higher average DMFT index, and poor maternal oral health has been associated with poor oral hygiene and ECC in their children.^23,26,27^

This present study found a positive, low magnitude association between child and mother salivary pH, which validates previous evidence.^18^ A Brazilian study showed in vitro that microcosm biofilm growth originating from mother-child pairs under regular sucrose exposure promotes the same cariogenic response independently of caries experience and microbiological profile, which possibly explains the relationship between maternal SSSpH and their child’s risk of developing ECC.^38^

This study’s findings validate previous evidence that child SSSpH is associated with ECC diagnosis and caries severity.^13,14,19^ Although findings to the contrary exist as Bowen^39^ argued that “Acid production is just one of many biological processes that occur within plaque when exposed to sugar” with salivary alkali production possibly impacting the development of ECC and Villavicencio found no association between children’s salivary pH and ECC.^40^

The strengths of this study include the large and geographically distributed sample size of ethnically diverse subjects which yielded over 80.0 percent post-hoc statistical power, with representation from three regions of the United States (i.e., Hawaii, New York, and Tennessee).

The study’s limitations include the lack of inter-rater reliability assessment for the resident-researchers; the potential that mother-child dyads were self-selected; low sensitivity (i.e., a high rate of false negatives); and over 80 percent of the sample with Medicaid as the payor, which serves as a proxy for low SES level. The high rate of false negatives may be attributed to confounding variables, which were not included in the study, such as the subject-specific salivary protective factors of buffering capacity, viscosity, flow rate, fluoride concentration; enamel strength, such as hardness, density, and fluorapatite concentration; and oral microbiome composition. The study consisted of mainly children from low SES families, who receive disproportionate oral health outcomes in comparison to their moderate and high SES counterparts,^41^ and so the results of this study may not be generalizable.

Future research may benefit from the inclusion of a larger and more SES diverse sample, additional subject-specific salivary protective factor variables, longitudinal design, and inter-rate reliability assessment to study the predictive strength of pH screening. Broader diversity of SES levels has the potential to improve analysis of salivary pH as a predictor for childhood ECC/SECC.

## CONCLUSIONS

Based upon the results of this study, the following conclusion can be made:

1. Dentists should understand that maternal SSSpH can signal the diagnosis and severity of early childhood caries.

## Data Availability

All data produced in the present study are available upon reasonable request to the authors.

## AKNOWLEDGMENTS

The authors acknowledge and thank the Hansjorg Wyss Department of Plastic Surgery, NYU Langone Health for funding support and pediatric dental residents Mohammad Allaoa, DMD, Tara Gainey, DDS, Johnny Joseph, DMD, Robert Kerns, DMD, MS, Spencer Kim, DDS, Kevin King, DDS, Jessica Kwon, DDS, Zohaib Munaf, DMD, MDS, Tashina Smiley, DMD, Yasmina Wright, DDS for data collection contributions.

## Conflict of Interests

The authors declare no conflict of interest.

## Notes

### Competing Interest Statement

The authors have declared no competing interest.

### Funding Statement

This study was funded by the Hansjorg Wyss Department of Plastic Surgery, NYU Langone Health.

### Author Declarations

Ethics committee/IRB of the NYU Grossman School of Medicine gave ethical approval for this work under protocol number 18-00076.

